# Sero-surveillance (IgG) of SARS-CoV-2 among Asymptomatic General population of Paschim Medinipur, West Bengal, India

**DOI:** 10.1101/2020.09.12.20193219

**Authors:** PS Satpati, SS Sarangi, KS Gantait, S Endow, NC Mandal, Kundu Panchanan, Bhunia Subhadip, Sarangi Soham

## Abstract

**Background:** Coronavirus disease 2019 (COVID-19) has emerged as a pandemic, and the infection due to SARSCoV-2 has now spread to more than 200 countries^3^. Surveillance systems form the foundation stone of active case finding, testing and contact tracing, which are the key components of the public health response to this novel, emerging infectious disease^4^. There is uncertainty about the true proportion of patients who remain asymptomatic or pre-symptomatic at a given time. As per the WHO-China Joint Monitoring Mission Report, and an analysis of 21 published reports, anywhere between 5 and 80 per cent of SARS-CoV-2-infected patients have been noted to be asymptomatic^5, 6^ Whereas in India 4197563 cases are positive, in which in West Bengal total 180788 cases(4.04% of Cases of India) positive of COVID 19. In Paschim Medinipur (West Medinipur) district contributing total 5489 cases (3.03% cases of West Bengal)^9,10,11^. In this scenario, we want to know the status of IgG seroprevalence of SARS-CoV-2 among asymptomatic general population, so that we can determine the extent of infection of SARS-CoV-2 in general population.

**Objectives:** <u>Primary Objective</u>:- To estimate the seroprevalence for SARS-CoV-2 infection in the general asymptomatic population at Paschim Medinipur District. <u>Secondary Objectives</u>-To estimate age and sex specific seroprevalence. To determine the socio demographic risk factors for SARS-CoV-2 infection; To determine the other risk factors like comorbidities, vaccination status, travel history, contact history etc.; To determine the durability of Immunity (IgG) conferred by natural infection of SARS-CoV-2 in individuals previously RTPCR positive.

**Methodology:** It was a cross sectional 30 cluster study among the population of Paschim Medinipur district of West Bengal conducted in last week of July and 1^st^ week of August 2020 among 458 asymptomatic general population and 30 RTPCR positive cases in 30 villages or wards of municipalities. 30 clusters were chosen from list of COVID 19 affected villages/wards of municipality as per PPS (Probability Proportional to Size) method.

**Results:** Of the 458 asymptomatic general population,19 asymptomatic people found to be seropositive IgG for SARS-CoV-2 with Mean or average total seropositivity rate of 4.15%. 19 Out of 30 (63.33%) RTPCR positive patients found Seronegative. Median of Days between RTPCR test and sero negativity found was 60 with minimum 28 days to maximum 101 days and Range of 73 days and a standard deviation of 19.46. Among risk factors, the risk of having IgG is more in persons having Travel history with odds ratio of 2.99-95%CI (1.17-7.65) with p-value-0.02. Hydroxychloroquine prophylaxis with Odds ratio of 8.49-95% CI(1.59-45.19) with p value - 0.003. Occupation as migrant labour with Odds ratio of 5.08-95% CI(1.96-13.18) with p value of 0.001. H/O Chicken pox with Odds ratio of 2.15-95% CI(0.59-7.79) with p value of 0.017. Influenza vaccinated with Odds ratio of 8.07 with 95% CI (0.8-81.48) with a p value of 0.036.

**Conclusion:** Of the 458 asymptomatic general population,19 asymptomatic people found to be seropositive IgG for SARS-CoV-2 with Mean or average total seropositivity rate of 4.15%. 19 Out of 30 (63.33%) RTPCR positive patients found Seronegative. Median of Days between RTPCR test and sero negativity found was 60 with minimum 28 days to maximum 101 days and Range of 73 days and a standard deviation of 19.46. Those having Travel History and having occupation as Migrant Labourer – have significantly higher probability of getting infected with SARS-CoV-2. No role has been found of Hydroxychloroquine Medicines as Chemoprophylactic. No durable immunity conferred by natural infection with SARS-CoV-2 –mean time to become seronegative after positive RTPCR test 60 days. So there is a chance of reinfection after average 2 months.

## INTRODUCTION

Serosurveys are studies that test body fluids, most commonly blood but also oral fluid, to estimate what proportion of the population has been vaccinated against or previously infected with a pathogen—and how many people remain susceptible^1^. Conducting population-based serosurveillance for severe acute respiratory syndrome-coronavirus-2 (SARS-CoV-2) will estimate and monitor the trend of infection in the adult general population, determine the socio-demographic risk factors and delineate the geographical spread of the infection^2^.

Coronavirus disease 2019 (COVID-19) has emerged as a pandemic, and the infection due to SARSCoV-2 has now spread to more than 200 countries^3^. As on 6^th^ September 2020, the Confirmed cases in India stood at 4197563, West Bengal had 180788 confirmed cases, and Paschim Medinipur District had 5489 confirmed cases.^9,10,11^ Surveillance systems form the foundation stone of active case finding, testing and contact tracing, which are the key components of the public health response to this novel, emerging infectious disease^4^. There is uncertainty about the true proportion of patients who remain asymptomatic or pre-symptomatic at a given time. As per the WHO-China Joint Monitoring Mission Report, and an analysis of 21 published reports, anywhere between 5 and 80 per cent of SARS-CoV-2-infected patients have been noted to be asymptomatic^5, 6^

The WHO global research map for COVID-19 and others recommend population-level sero epidemiological studies to generate data on the levels of infection in populations and recommend containment measures accordingly^7^. In order to achieve this, a protocol for conducting such population-based sero-epidemiological investigation for COVID-19 has been proposed by the WHO^8^.

**Serosurveys** involve collection of specimens to measure the presence and level of antigen-specific antibodies in a group of people. It reveals what proportion of the population has been exposed to an infectious disease or has been vaccinated against a pathogen. It also estimates the level of population immunity to one or more infectious diseases; Sero Survey identifies gaps in immunity because people were not vaccinated or previously infected. Immunity gaps can be in specific age groups, certain locations, or among specific populations such as migrants. Identifying immunity gaps can guide immunization programs. It estimates parameters for modelling and transmission dynamics to measure disease burden and guide immunization programs^12^. Serologic studies are crucial for clarifying dynamics of the coronavirus disease pandemic. Past work on serological studies (e.g., during influenza pandemics) has made relevant contributions. Although detection of antibodies to measure exposure, immunity, or both seems straightforward conceptually, numerous challenges exist in terms of sample collection, what the presence of antibodies means, and appropriate analysis and interpretation to account for test accuracy and sampling biases. Successful deployment of serologic studies depends on type and performance of serologic tests, population studies, use of adequate study designs, and appropriate analysis and interpretation of data. We highlight key questions that serologic studies can help answer at different times, review strengths and limitations of different assay types and study designs, and discuss methods for rapid sharing and analysis of serologic data to determine the extent of transmission of SARS-CoV-2. SARS-CoV-2 serologic studies has largely focused on 2 questions: first, what proportion of a population has been infected; and second, what proportion of a population is immune to disease or infection? Firstly, for infections that elicit detectable antibody responses, serologic studies can detect past infection regardless of clinical symptoms. This capability is useful for understanding the extent of past transmission^13^. Secondly, if measured antibody responses correlate with protection, serologic studies can be used to measure the proportion of the population those who are immune. This information can be used to guide control policies, help identify populations that are still susceptible to epidemics, target treatment or vaccination trials, and target vaccination when available. Although much discussion around use of serologic testing to inform persons of their serologic status has occurred, crucial distinctions exist between the use of serologic information to estimate population-level versus person-level immunity. Person-level immunity information is currently fraught with scientific, ethical, and legal uncertainties, which we do not address in this article.

## OBJECTIVES

### Primary Objective

To estimate the seroprevalence for SARS-CoV-2 infection in the general asymptomatic population at District level.

### Secondary Objectives

(i) To estimate age and sex specific seroprevalence; (ii) To determine the socio demographic risk factors for SARS-CoV-2 infection; (iii) To determine the other risk factors like comorbidities, vaccination status, travel history, contact history etc; (iv) To determine the durability of Immunity (IgG) conferred by natural infection of SARS-CoV-2 in individuals previously RTPCR positive.

## METHODOLOGY

Study design—Cross sectional 30 Cluster study

Study Population-

- Total Population of Paschim Medinipur District-52,40,571
- Total Number of affected Block/Municipality-28
- Total Number of CoVID 19 affected villages/wards-219

### For case definitions

We have followed the WHO^1^ and ICMR^2^ Protocols for Serosurvey

### Sampling method-30 cluster study

30 clusters were chosen from list of COVID 19 affected villages/wards of municipality as per PPS (Proportionate to population size) method.

### Total Sample Size

450 as calculated by StatCalc of Epi-info 7. So, 15 samples per 30 clusters were tested. Finally, 458 Samples were collected from the selected 30 Clusters. Along with these, 30 Samples of previously RT-PCR positive individuals were collected.

### Ethical Clearance

was taken from Medinipur Medical College Ethical Clearance Committee.

### Method of data collection

30 (thirty), two membered team comprising of concerned PHN and BPHN, MT (Lab) detailed from same block visited the selected village and PHN/BPHN collected data as per pre-tested Semi structured schedule and MT drew blood and after making serum, transport it as per protocol maintaining cold chain to VRDL Lab of MMCH.

### Lab Test

IgG ELISA was done using ErbaLisa COVID-19 IgG ELISA Kit [Manufactured by Calbiotech Inc., USA (Erba-TransAsia Group Company)] having a Sensitivity of 98.3% and Specificity of 98.1%.

### Data Analysis

Data Analysis was done by SPSS 27 and Epi Info 7. For Sociodemographic Variables and Risk Factors, comparison done between IgG positive and IgG Negative Groups-Odds Ratio calculated (95% Interval), Chi-Square value, degree of freedom and p-value (<0.05) calculated and factors found with strong association and significant p value put in Multinomial Logistic Regression Analysis.

## RESULTS

A 30 cluster cross-sectional study conducted in the last week of July and 1^st^ week of August 2020 among 458 asymptomatic general population and 30 RTPCR positive cases in 30 villages or wards of municipalities.

### Descriptive Epidemiology of the Asymptomatic General Population (Table 1)

**Table 1:**
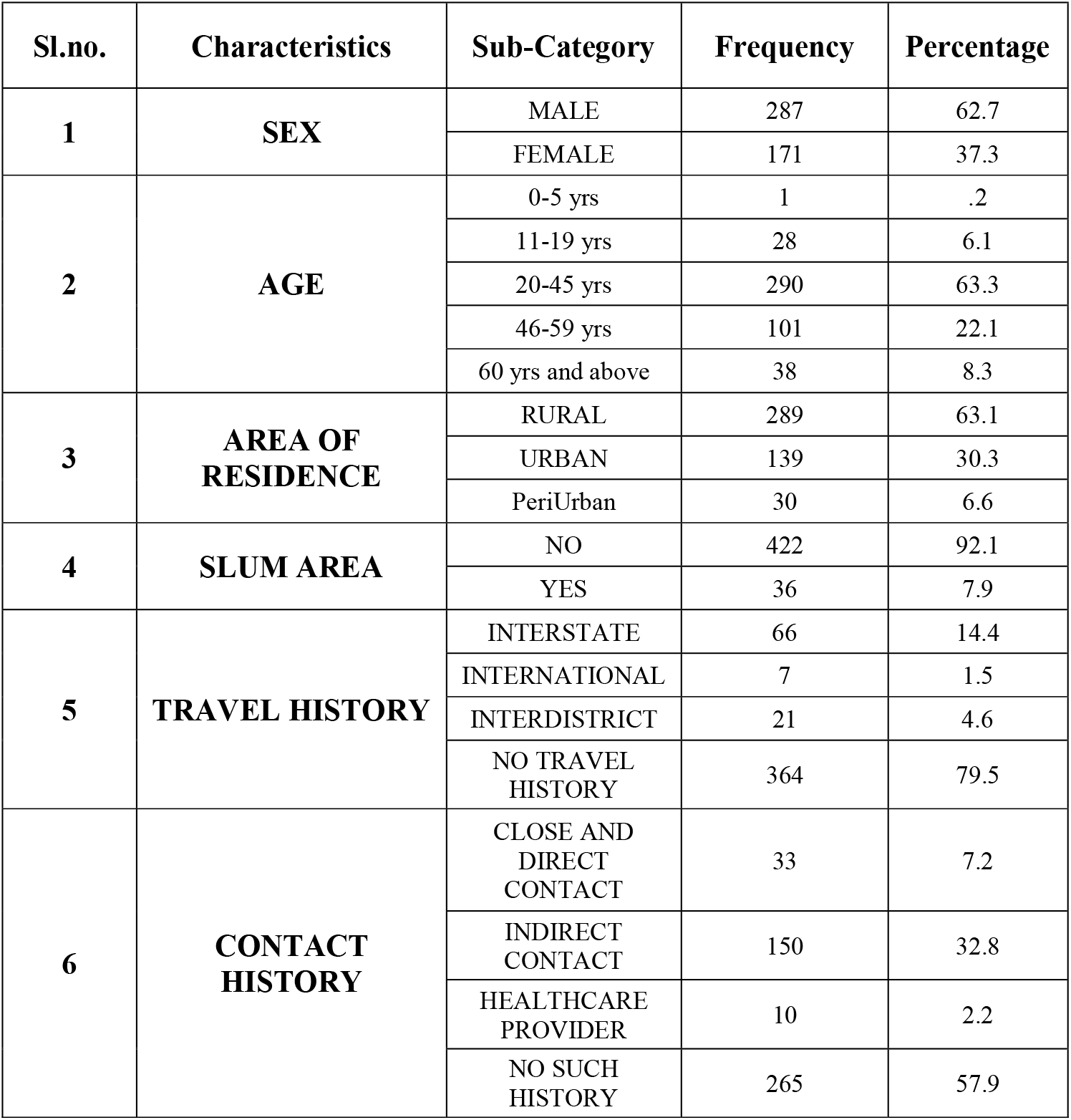
Descriptive Epidemiology.

Among the samples collected, 62.7% were Males (n = 287), and 37.3% were Females (n = 171). The highest no. of samples were from 20-45 years of age group (63.3%; n = 290). 63.1% persons were from Rural Area (n = 289), 30.3% persons were from Urban Area (n = 139). 6.6% of the persons lived in a Periurban Area (n = 30). 7.9% Samples were from Slum Area (n = 36). 20.5% Persons had History of Travel(n = 94) [Interstate-14.4%; International-1.5%; Interdistrict-4.6%]. 79.5% Persons had no History of Travel. 42.1% of the persons had Contact History with a Covid-19 Patient (n = 193) [Close and Direct Contact-7.2%; Indirect Contact-32.8%; Healthcare Provider-2.2%].

### IgG Serosurvey Results of Pachim Medinipur District among Asymptomatic General Population of Paschim Medinipur District (Table 2)

**Table 2:** SARS-CoV-2 SERO-SURVEILLANCE RESULTS IN GENERAL POPULATION OF PASCHIM MEDINIPUR.

| BLOCK | Total no. of samples collected for Serosurveillance | No. of IgG Positive | No. of Negative IgG | Seropositivity Percentage (95% CI) |
| --- | --- | --- | --- | --- |
| Midnapore Municipality | 16 | 1 | 15 | 6.25(0.16-30.23) |
| Ghatal Municipality | 16 | 2 | 14 | 12.50(1.55-38.35) |
| Garhbeta I Block | 15 | 0 | 15 | 0.00 |
| Chandrakona Municipality | 15 | 0 | 15 | 0.00 |
| Dantan II | 15 | 0 | 15 | 0.00 |
| Dantan I | 14 | 0 | 14 | 0.00 |
| KGP Municipality | 108 | 4 | 104 | 3.70(1.02-9.21) |
| Daspur I | 75 | 3 | 72 | 4.00(0.83-11.25) |
| Daspur II | 92 | 9 | 83 | 9.78(4.57-17.76) |
| Ghatal Block | 30 | 0 | 30 | 0.00 |
| Chandrakona I | 15 | 0 | 15 | 0.00 |
| Pingla | 16 | 0 | 16 | 0.00 |
| Keshpur | 31 | 0 | 31 | 0.00 |
| **TOTAL** | **458** | **19** | **439** | **4.15 (2.52-6.40)** |

19 asymptomatic persons found to be seropositive IgG for SARS-CoV-2 with Mean or average total seropositivity rate of 4.15% [95% Confidence Interval-Lower-2.52% Upper-6.40%]. 439 asymptomatic general persons were found to be Seronegative(95.85%). Highest Seropositivity percentage found in Ghatal Municipality of 12.50% followed by Daspur II of 9.78%, Daspur I of 4.00% and Kharagpur Municipality of 3.70% and Midnapur Municipality of 6.25%.

### Socio-demographic Factors associated with Increased Risk of IgG Seropostivity of COVD-19 (Table-3) were

**Table 3:** SOCIO-DEMOGRAPHIC FACTORS.

| Sl. No | CHARACTERISTICS | SUB-CATEGORY | IgG Negative n(%) | IgG Positive n(%) | Odds Ratio | 95% Conf. Interval of Odds Ratio |  | chi-square | p—value (Significant for p<0.05) |
| --- | --- | --- | --- | --- | --- | --- | --- | --- | --- |
|  |  |  |  |  |  | Lower | Upper |  |  |
| 1 | SEX | MALE (n=287) | 272(94.77%) | 15(5.23%) | 2.3 | 0.75 | 7.05 | 2.24 | 0.13 |
|  |  | FEMALE (n=171) | 167(97.66%) | 4(2.34%) |  |  |  |  |  |
| 2 | AGE-GROUP | 0-5 yrs (n=1) | 1(100.00%) | 0(0.00%) |  |  |  |  |  |
|  |  | 11-19 yrs (n=28) | 25(89.29%) | 3(10.71%) |  |  |  |  |  |
|  |  | 20-45 yrs (n=290) | 279(96.21%) | 11(3.79%) | 3.02 | 0.82 | 11.11 | 3.022 | 0.082 |
|  |  | 46-59 yrs (n=101) | 97(96.04%) | 4(3.96%) |  |  |  |  |  |
|  |  | 60 yrs & above (n=38) | 37(97.37%) | 1(2.63%) |  |  |  |  |  |
| 3 | LOCALITY | RURAL(n=289) | 275(95.16%) | 14(4.84%) | 1.67 | 0.59 | 4.72 | 0.95 | 0.33 |
|  |  | URBAN(n=139) | 134(96.40%) | 5(3.60%) | 1.23 | 0.43 | 3.48 | 0.15 | 0.696 |
|  |  | PeriUrban(n=30) | 30(100.00%) | 0(0.00%) |  |  |  |  |  |
| 4 | SLUM AREA | NO (n=422) | 406(96.21%) | 16(3.79%) |  |  |  |  |  |
|  |  | YES (n=36) | 33(91.67%) | 3(8.33%) | 2.30 | 0.64 | 8.32 | 1.72 | 0.19 |
| 5 | EMPLOYMENT | MIGRANT LABOUR (n=63) | 55(87.30%) | 8(12.70%) | 5.07 | 1.96 | 13.18 | 13.42 | 0.00 |
|  |  | GOVT EMPLOYEE (n=47) | 45(95.74%) | 2(4.26%) |  |  |  |  |  |
|  |  | PRIVATE EMPLOYEE (n=19) | 18(94.74%) | 1(5.26%) |  |  |  |  |  |
|  |  | HEALTHCARE PROVIDER (n=18) | 17(94.44%) | 1(5.56%) |  |  |  |  |  |
|  |  | AGRICULTURE (n=53) | 53(100.00%) | 0(0.00%) |  |  |  |  |  |
|  |  | BUSINESS (n=43) | 42(97.67%) | 1(2.33%) |  |  |  |  |  |
|  |  | OTHERS (n=168) | 163(97.02%) | 5(2.98%) |  |  |  |  |  |
|  |  | UNEMPLOYED (n=47) | 46(97.87%) | 1(2.13%) |  |  |  |  |  |
| 6 | Educational Qualification | ILLITERATE (n=40) | 39(97.5%) | 1(2.5%) |  |  |  |  |  |
|  |  | PRE-PRIMARY (n=16) | 16(100.00%) | 0(0.00%) |  |  |  |  |  |
|  |  | PRIMARY (n=163) | 157(96.32%) | 6(3.68%) | 0.82 | 0.31 | 2.22 | 0.14 | 0.71 |
|  |  | SECONDARY (n=100) | 93(93.00%) | 7(7.00%) |  |  |  |  |  |
|  |  | HIGHER SECONDARY (n=52) | 51(98.08%) | 1(1.92%) |  |  |  |  |  |
|  |  | GRADUATION (n=64) | 61(95.31%) | 3(4.69%) |  |  |  |  |  |
|  |  | POST GRADUATION (n=23) | 22(95.65%) | 1(4.35%) |  |  |  |  |  |
| 7 | Monthly Income | UPTO RS1000 (n=93) | 91(97.85%) | 2(2.15%) |  |  |  |  |  |
|  |  | 1001-3000 (n=43) | 43(100.00%) | 0(0.00%) |  |  |  |  |  |
|  |  | 3001-5000 (n=103) | 98(95.15%) | 5(4.85%) | 1.24 | 0.44 | 3.53 | 0.17 | 0.68 |
|  |  | 5001-10000 (n=116) | 110(94.83%) | 6(5.17%) | 1.38 | 0.51 | 3.72 | 0.41 | 0.52 |
|  |  | 10001-25000 (n=50) | 46(92.00%) | 4(8.00%) |  |  |  |  |  |
|  |  | 25001-50000 (n=32) | 31(96.88%) | 1(3.12%) |  |  |  |  |  |
|  |  | ABOVE 50000 (n=21) | 20(95.24%) | 1(4.76%) |  |  |  |  |  |

Male Sex – with odds ratio of 2.3 (95% conf. Int.-0.75<OR< 7.05); Age group 20-45 Years - with odds ratio 3.02 (95% conf. Int.-0.82<OR< 11.11); Inhabitant of Slum Area-with Odds Ratio of 2.30 (95% conf. Int.-0.64<OR< 8.32); Migrant Labour as Occupation – with Odds Ratio of 5.07 (95% conf. Int.-1.96<OR< 13.18).

Though the above factors had high odds ratio, only Migrant Labour as Occupation was found as a statistically significant Socio-Demographic Risk Factor for Covid-19 Seropositivity (p = 0.001). All other socio-demographic factors had p>0.05. Also, IgG Seropositivity was highest among Migrant Labour group – 12.70%.

No Comorbidities were found significantly associated with increased risk of IgG Seropositivity (Comorbidity Profile - Table 4)

**Table 4:** Comorbidity Profile.

| SL. NO | COMORBIDITIES | SUB-CATEGORY | IgG Negative n(%) | IgG Positive n(%) | Odds Ratio | 95% Confidence Limit for Odd's Ratio |  | chi—square | df | p—value (Significant for p<0.05) |
| --- | --- | --- | --- | --- | --- | --- | --- | --- | --- | --- |
|  |  |  |  |  |  | Upper | Lower |  |  |  |
| **1** | **DIABETES** | NO (n=418) | 400 (95.69%) | 18 (4.31%) |  |  |  |  |  |  |
|  |  | **YES (n=40)** | **39 (97.5%)** | **1 (2.5%)** | **0.57** | **0.07** | **4.38** | **0.299** | **1** | **0.584** |
| **2** | **DIABETES MEDICATION TAKEN** | NO (n=425) | 407 (95.76%) | 18 (4.24%) |  |  |  |  |  |  |
|  |  | **YES (n=33)** | **32 (96.97%)** | **1 (3.03%)** | **0.71** | **0.03** | **4.08** | **0.11** | **1** | **0.74** |
| **3** | **BLOOD SUGAR (RANDOM) (IN mg/dl)** | **MORE THAN 140 (n=51)** | **50 (98.04%)** | **1 (1.96%)** | **0.43** | **0.05** | **3.3** | **0.69** | **1** | **0.405** |
|  |  | LESS THAN 140 (n=407) | 389 (95.58%) | 18 (4.42%) |  |  |  |  |  |  |
| **4** | **HYPERTENSION** | NO (n=405) | 387 (95.56%) | 18 (4.44%) |  |  |  |  |  |  |
|  |  | **YES (n=53)** | **52 (98.11%)** | **1 (1.89%)** | **0.41** | **0.05** | **3.16** | **0.77** | **1** | **0.38** |
| **5** | **ANTI-HYPERTENSIVE DRUGS TAKEN** | NO (n=417) | 399 (95.68%) | 18 (4.31%) |  |  |  |  |  |  |
|  |  | **YES (n=41)** | **40 (97.56%)** | **1 (2.44%)** | **0.55** | **0.07** | **4.26** | **0.33** | **1** | **0.57** |

### Risk factors associated statistically significantly with IgG Seropositivity of COVID-19 (p<0.05) (Table-5)

**Table 5:** Summary of Risk FACTORS for SEROPOSITIVITY.

| SL. NO | RISK FACTORS | SUB-CATEGORY | IgG Negative n(%) | IgG Positive n(%) | Odds Ratio | 95% Confidence Limit for Odd's Ratio |  | chi—square value | df | p—value (Significant for p<0.05) |
| --- | --- | --- | --- | --- | --- | --- | --- | --- | --- | --- |
|  |  |  |  |  |  | U p p e r | Lo wer |  |  |  |
| 1 | TRAVEL HISTORY | NO TRAVEL HISTORY (n=364) | 353(96.98%) | 11 (3.02%) |  |  |  |  |  |  |
|  |  | TRAVEL HISTORY PRESENT (n=94) | 86(91.49%) | 8(8.51%) | 2.99 | 1.17 | 7.65 | 5.66 | 1 | 0.02 |
| 2 | HYDROXYCHLOROQUINE PROPHYLAXIS | NO(n=450) | 433(96.22%) | 17(3.78%) |  |  |  |  |  |  |
|  |  | YES (n=8) | 6(75.0%) | 2(25.0%) | 8.49 | 1.59 | 45.19 | 8.9 | 1 | 0.003 |
| 3 | MIGRANT LABOURER | NOT A MIGRANT (n=395) | 384(97.21%) | 11(2.78%) |  |  |  |  |  |  |
|  |  | MIGRANT LABOURER (n=63) | 55(87.30%) | 8(12.70%) | 5.08 | 1.96 | 13.18 | 13.43 | 1 | 0.001 |
| 4 | HISTORY OF CHICKEN POX | NO (n=270) | 264(97.78%) | 6(2.22%) |  |  |  |  |  |  |
|  |  | UNKNOWN (n=102) | 93(91.18%) | 9(8.82%) |  |  |  |  |  |  |
|  |  | YES (n=86) | 82(95.34%) | 4(4.65%) | 2.15 | 0.59 | 7.79 | 8.18 | 2 | 0.017 |
| 5 | INFLUENZA VACCINATION STATUS | None (n=235) | 228(97.02%) | 7(2.98%) |  |  |  |  |  |  |
|  |  | Unknown (n=219) | 208(94.98%) | 11(5.02%) |  |  |  |  |  |  |
|  |  | Yes (n=4) | 3(75.0%) | 1(25.0%) | 8.07 | 0.8 | 81.48 | 4.41 | 1 | 0.036 |

**(i) Travel history with odds ratio of 2.99-** 95%CI (1.17-7.65) with **p-value-0.02**;(ii) **Hydroxychloroquine prophylaxis** with **Odds ratio of 8.49-** 95% CI(1.59-45.19) with **p value - 0.003; (iii) Occupation as Migrant Labour** with **Odds ratio of 5.08-** 95% CI(1.96-13.18) with **p value of 0.001;(iv) H/O Chicken pox** with **Odds ratio of 2.15-** 95% CI(0.59-7.79) with **p value of 0.017;(v) Influenza vaccinated** with **Odds ratio of 8.07** with 95% CI (0.8-81.48) with a **p value of 0.036**.

Finally, **Multinomial Logistic Regression was done (Table-6):-** Through Multinomial Logistic Regression Model having the above associated risk factors from Table 8 as Independent Variable and IgG Seropositivity as Dependent Variable as these Independent Variable had High Odds ratio and significant p-value (p<0.05). The Regression Model had a Model Fitting Significance of 0.005 and therefore was significant. From the Regression model, it was found that –

- Occupation as Migrant Labour was significant with Significance of 0.038 and Exp.(B) of 8.007
- Hydroxychloroquine Intake was significant with Significance of 0.045 and Exp.(B) of 7.290

**Table 6:** Multinomial Logistic Regression Table.

**Model Fitting Information**
| Model | Model Fitting Criteria | Likelihood Ratio Tests |  |  |
| --- | --- | --- | --- | --- |
|  | -2 Log Likelihood | Chi-Square | df | Sig. |
| Intercept Only | 58.030 |  |  |  |
| Final | 37.494 | 20.535 | 7 | .005 |

**Parameter Estimates**
| IgG Status OR Covid 19 Seropositivity |  | B | Std. Error | Wald | df | Sig. | Exp (B) | 95% Confidence Interval for Exp(B) |  |
| --- | --- | --- | --- | --- | --- | --- | --- | --- | --- |
|  |  |  |  |  |  |  |  | Lower Bound | Upper Bound |
| **POSITIVE** | Intercept | -2.412 | 1.614 | 2.232 | 1 | .135 |  |  |  |
|  | [occupation_code=.00] (Migrant Labourer) | 2.080 | 1.003 | 4.306 | 1 | .038 | 8.00 | 1.122 | 57.131 |
|  | [occupation_code=1.00] (Not a Migrant Labourer) | 0<sup>b</sup> |  |  | 0 |  |  |  |  |
|  | [HYDROXYCHLOROQUINE TAKEN=0] (Yes) | 1.986 | .992 | 4.008 | 1 | .045 | 7.29 | 1.043 | 50.971 |
|  | [HYDROXYCHLOROQUINE TAKEN=1] (No) | 0<sup>b</sup> |  |  | 0 |  |  |  |  |
|  | [Travel_History_recode=0] (No Travel History) | .785 | 1.019 | .594 | 1 | .441 | 2.19 | .297 | 16.164 |
|  | [Travel_History_recode=1] (Travel History Present) | 0<sup>b</sup> |  |  | 0 |  |  |  |  |
|  | [chickenpox_hist_final=0] (No) | -.447 | .776 | .331 | 1 | .565 | .640 | .140 | 2.931 |
|  | [chickenpox_hist_final=1] (Unknown) | .868 | .915 | .900 | 1 | .343 | 2.38 | .396 | 14.315 |
|  | [chickenpox_hist_final=2] (Yes) | 0<sup>b</sup> |  |  | 0 |  |  |  |  |
|  | [influenza_vacn_final=0] (No) | -1.962 | 1.468 | 1.787 | 1 | .181 | .141 | .008 | 2.495 |
|  | [influenza_vacn_final=1] (Unknown) | -2.118 | 1.530 | 1.916 | 1 | .166 | .120 | .006 | 2.414 |
|  | [influenza_vacn_final=2] (Yes) | 0<sup>b</sup> |  |  | 0 |  |  |  |  |

### IgG Serosurvey Results of Pachim Medinipur District among Previously RT-PCR Positive of Paschim Medinipur District(Table 7, Table 8)

**Table 7:** SERO-SURVEY RESULTS AMONG PREVIOUSLY COVID-19 RTPCR POSITIVE PATIENTS IN PASCHIM MEDINIPUR.

| BLOCK | Total no. of samples collected for Serosurveillance | No. of IgG Positive | No. of IgG Negative | Seropositivity Percentage |
| --- | --- | --- | --- | --- |
| Ghatal Municipality | 1 | 0 | 1 | 0.00 |
| Garhbeta I Block | 1 | 0 | 1 | 0.00 |
| Chandrakona Municipality | 1 | 0 | 1 | 0.00 |
| Dantan II | 1 | 0 | 1 | 0.00 |
| KGP Municipality | 7 | 4 | 3 | 57.14 |
| Daspur I | 8 | 2 | 6 | 25.00 |
| Daspur II | 8 | 5 | 3 | 62.50 |
| Ghatal Block | 1 | 0 | 1 | 0.00 |
| Chandrakona I | 1 | 0 | 1 | 0.00 |
| Keshpur | 1 | 0 | 1 | 0.00 |
| **TOTAL** | **30** | **11** | **19** | **36.67** |

**Table 8:** RELATIONSHIP OF “DAYS BETWEEN IgG ELISA TEST & RTPCR TEST” AND IgG SERO-STATUS.

| Statistics |  |  |  |
| --- | --- | --- | --- |
| Days between IgG and rtPCR |  |  |  |
| IgG NEGATIVE | N | Valid | 19 |
|  |  | Missing | 0 |
|  | Mean |  | 60.32 |
|  | Std. Error of Mean |  | 4.464 |
|  | Median |  | 60.00 |
|  | Mode |  | 49<sup>a</sup> |
|  | Std. Deviation |  | 19.460 |
|  | Range |  | 73 |
|  | Minimum |  | 28 |
|  | Maximum |  | 101 |
| IgG POSITIVE | N | Valid | 11 |
|  |  | Missing | 0 |
|  | Mean |  | 48.09 |
|  | Std. Error of Mean |  | 4.216 |
|  | Median |  | 49.00 |
|  | Mode |  | 31<sup>a</sup> |
|  | Std. Deviation |  | 13.982 |
|  | Range |  | 36 |
|  | Minimum |  | 31 |
|  | Maximum |  | 67 |
| a. Multiple modes exist. The smallest value is shown |  |  |  |

Out of 30 Samples Collected, only 11 previously RT-PCR Positive Individuals were found IgG Seropositive for COVID-19 (36.67%). 19 Persons were found Seronegative (63.33%)[Table 7]. The Mean and Median no. of Days Between RTPCR test and IgG Test in IgG Negative Persons were 60.32 and 60 respectively with a minimum of 28 days and Maximum of 101 Days. (Table 8). Whereas, in the case of IgG Positive Individuals, the Mean and Median No. of Days between RTPCR and IgG ELISA Test were 48.09 and 49 respectively with a minimum of 31 days and Maximum of 67 days (Table 8).

## DISCUSSION

The findings of the IgG Serosurvey conducted during last week of July to first week of August indicated that 4.15 percent of the asymptomatic General Population of Paschim Medinipur District, West Bengal, India were exposed to SARS-CoV-2 infection. The seroprevalence ranged from 12.50% in Ghatal Municipality to 0.00 % in Garhbeta-I, Dantan-I, Dantan-II, Chandrakona-I, Chandrakona Municipality, Ghatal Block, Pingla and Keshpur blocks of the district.

As per the Nationwide Serosurvey conducted by ICMR in May-June 2020^33^, the nationwide seroprevalence was 0.73 percent. Though our seroprevalence was higher than the national average, it was much lower than the 22.86% seroprevalence in Delhi. Since the seroprevalence in our district is still low, it indicates that a majority of the population of our district was still susceptible to COVID-19 infection. It also correlates with the study done in England in which they have found around 4.2% sero positivity in the month of May 2020^15^. A dashboard of sero-epidemiological data available from 22 countries estimated the pooled seroprevalence to be 4.76 per cent, ranging from 0.65 percent in Scotland to 26.6 per cent in Iran^36^. Other studies also correlate the same prevalence in the young population in different countries^20,21,22,23,24,25^ In this study in 20 to 45 yr age group we have found that 3.79% IgG positivity which is around same percentage which in the seroprevalence study of Switzerland^14^ found in week 1 around first week of April 2020 found (around 3.5%). Seropositivity was higher among Males (5.23%). This is also seen in the ICMR Serosurvey^33^, and other studies around the globe.^20,21,22,23,24,25^. In India, Migrant Labours have been regarded as a high risk group for SARS-CoV-2 infection. This is also reflected by the fact that being a Migrant Labour was significantly associated with IgG Seropositivity with Odds Ratio of 5.07 (95% conf. Interval-1.96<OR<13.18) with p-value of 0.00 (Significant since p<0.05).

Another significant finding of our study was the strong association of Hydroxychloroquine Intake with COVID-19 Seropositivity. This shows the ineffectiveness of Hydroxychloroquine in preventing COVID-19 infection. Not only that, high odds ratio with significant p-value also indicates that those who had taken Hydroxychloroquine had very high risk of COVID-19 infection. This requires further research. Though, trials of Hydroxychloroquine as a post-exposure prophylaxis for COVID-19 in USA^34^ and UK^35^ have already found it ineffective.

Other risk factors that were significantly associated with seropositivity were-Travel History, History of Chicken Pox and History of Influenza Vaccination. Though having travel history increases risk of exposure to a COVID-19 positive patient, other factors require further research to establish definitive association between these factors and COVID-19 seropositivity.

We have not found any correlation between Diabetes or persons who were taking medication for diabetes and IgG positivity in this study. Also, no correlation was found between Hypertension and IgG Positivity. This differs from a South Korean study(Song SK et. al.)^31^, which found Diabetes as a risk factor for IgG Seropositivity.

In our study, we found that only 36.67 percent of individuals who previously were RT PCR positive for COVD-19, had IgG antibodies. This is in correlation with the Delhi Serosurvey^37^, which found that only 53.37% of individuals who previously were RT PCR positive for COVD-19, had IgG antibodies. In the study of Japan –“Of the139 COVID-19 serum specimens, IgM was detected in 27.8 %, 48.0 %, and 95.8 % of the specimens collected within 1 week, 1-2 weeks, and > 2 weeks after symptom onset and IgG was detected in 3.3 %, 8.0 %, and 62.5 %, respectively”^32^. This study also correlates with our study. This indicates that previous infection with SARS-CoV-2 does not provide long-term humoral immunity as determined by IgG Seropositivity. There is thus, a chance of reinfection after recovering from COVID-19.

## CONCLUSION

Cross-sectional 30 cluster study was conducted in the district of Paschim Medinipur, West Bengal, India during last week of July and 1^st^ week of August 2020 as a joint venture of VRDL lab MMCH and Health and Family Welfare Department, Paschim Medinipur, Government of West Bengal, India. Seropositivity (IgG) rate among asymptomatic general population overall in Paschim Medinipur district is 4.15% (95% CI-2.52%-6.40%) with highest in Ghatal Municipality 12.5% and Daspur II block of 9.78%. Those having Travel History, and having occupation as Migrant Labourer – have significantly higher probability of getting infected with SARS-CoV-2. In patients who had taken Hydroxy Chloroquine as a chemoprophylactic, the probability of getting infected with SARS-CoV-2 was significantly higher. No role has been found for Hydroxychloroquine as Chemoprophylactic. No durable immunity conferred by natural infection with SARS COV2 –mean time to become seronegative after positive RTPCR test 60 days. So, there is a chance of reinfection after 2 months. Those who were vaccinated by influenza vaccine previously and H/O chicken pox are more prone to become IgG Seropositive of SARS COV2 among asymptomatic healthy populations.

## Data Availability

All the data referred to in the manuscript is available with the authors and this includes the relevant data of the subjects (Collected through Google Forms), their consent forms, and all the Excel Sheets and SPSS Statistics Files used for data entry and analysis. If requested, those may be submitted.

## Acknowledgements

We acknowledge the contributions and support of all staffs and officers of Department of Health and Family Welfare, Paschim Medinipur and all scientists, staffs, Laboratory Technicians, Data Entry Operators engaged in VRDL Lab (ICMR), Midnapore Medical College and Hospital, Paschim Medinipur.

